# Rheumatoid arthritis synovial fluid neutrophils drive inflammation through production of chemokines, reactive oxygen species and neutrophil extracellular traps

**DOI:** 10.1101/2020.07.16.20155291

**Authors:** Helen L Wright, Max Lyon, Elinor A Chapman, Robert J Moots, Steven W Edwards

## Abstract

Rheumatoid arthritis (RA) is a chronic inflammatory disorder affecting synovial joints. Neutrophils are believed to play an important role in both the initiation and progression of RA, and large numbers of activated neutrophils are found within both synovial fluid (SF) and synovial tissue from RA joints. In this study we analysed paired blood and SF neutrophils from patients with severe, active RA (DAS28> 5.1, n=3) using RNA-seq. 772 genes were significantly different between blood and SF neutrophils. IPA analysis predicted that SF neutrophils had increased expression of chemokines and ROS production, delayed apoptosis, and activation of signalling cascades regulating the production of NETs. This activated phenotype was confirmed experimentally by incubating healthy control neutrophils in cell-free RA SF, which was able to delay apoptosis and induce ROS production in both unprimed and TNF*α* primed neutrophils (p< 0.05). RA SF significantly increased neutrophil migration through 3mM transwell chambers (p< 0.05) and also increased production of NETs by healthy control neutrophils, including exposure of myeloperoxidase (MPO) and citrullinated histone-H3-positive DNA NETs. IPA analysis predicted NET production was mediated by signalling networks including AKT, RAF1, SRC and NF-*κ*B. Our results expand the understanding of the molecular changes that take place in the neutrophil transcriptome during migration into inflamed joints in RA, and the altered phenotype in RA SF neutrophils. Specifically, RA SF neutrophils lose their migratory properties, residing within the joint to generate signals that promote joint damage, as well as inflammation via recruitment and activation of both innate and adaptive immune cells. We propose that this activated SF neutrophil phenotype contributes to the chronic inflammation and progressive damage to cartilage and bone observed in patients with RA.

## 1 INTRODUCTION

Rheumatoid arthritis (RA) is an inflammatory disorder characterised by systemic inflammation, including swelling and pain in synovial joints. Left untreated, uncontrolled inflammation will destroy joints, causing deformity and disability. Many studies have shown that blood neutrophils have an aberrant, activated phenotype in RA, characterised by increased production of ROS and cytokines, and delayed apoptosis (Wright et al., 2014b, 2011; Martelli-Palomino et al., 2017; Marchi et al., 2018). As well as having an activated phenotype in peripheral blood, activated neutrophils are found at high numbers in both synovial joints and tissues of patients with RA (Wittkowski et al., 2007; Turunen et al., 2016; Mourao et al., 2010). Their presence is accompanied by high levels of neutrophil granule proteins in synovial fluid, including myeloperoxidase (MPO), cathepsin G, proteinase 3, elastase and lactoferrin (Wright et al., 2014b; Edwards et al., 1988; Thieblemont et al., 2016; Wong et al., 2009; Nzeusseu Toukap et al., 2014). These granule proteins contribute to the pathogenesis of RA through proteolytic cleavage and activation of proteins (including cytokines and chemokines), cleavage of soluble receptors to initiate trans-signalling (such as the IL-6 receptor) and degradation of cartilage (e.g. cleavage of collagen fibres) (Wright et al., 2014b; Pham, 2006; Baici et al., 1982; Van den Steen et al., 2002; Desgeorges et al., 1997). *Ex vivo* synovial fluid (SF) neutrophils have an altered phenotype compared to paired blood neutrophils for example displaying higher levels of superoxide (O_2_^•-^) production, and containing phosphorylated p47^phox^, indicating assembly and activation of the NADPH oxidase (NOX2) *in vivo* (El Benna et al., 2002). They also express the high-affinity Fc*γ*R1 receptor (CD64) and MHC Class II proteins (Cross et al., 2003; Quayle et al., 1997; Robinson et al., 1992; Cedergren et al., 2007). SF neutrophil lysates also have lower levels of granule proteins such as MPO, confirming their degranulation within the synovial joint (Edwards et al., 1988). Animal studies and human case studies of early RA suggest an important role for neutrophils in the initiation of synovial inflammation in RA joints (Wittkowski et al., 2007; Mourao et al., 2010; Wipke and Allen, 2001), possibly through the release of granule enzymes and production of VEGF, both of which enable fibroblast adhesion and growth of the inflammatory synovial pannus (Kasama et al., 2000; McCurdy et al., 1995). A key role for exposure of citrullinated antigens on neutrophil extracellular traps (NETs) has been proposed in the initiation of auto-immunity and development of anti-citrullinated peptide auto-antibodies (ACPA) in RA (Wright et al., 2014b; Khandpur et al., 2013). NET products are present in both RA serum and synovial fluid (Pieterse et al., 2018) and have also been observed in synovial biopsy tissues from RA patients, characterised by positive staining for CD15, elastase, MPO and citrullinated (cit) histone H3 (Khandpur et al., 2013; Spengler et al., 2015). It was recently shown that up to 70% of newly-diagnosed RA patients have auto-antibodies in their serum that recognise NET components (ANETA) (de Bont et al., 2020).

We have extensively studied neutrophil phenotype in RA and shown that RA neutrophils have activated NF-*κ*B signalling leading to delayed apoptosis (Wright et al., 2011). Additionally, we have shown using RNA-seq that neutrophil gene expression is altered in RA compared to healthy controls (Wright et al., 2015). Gene expression in RA patients pre-TNFi therapy can be used to stratify patients based on response or non-response to therapy (Wright et al., 2017). Whilst several studies have analysed SF neutrophil functions, to our knowledge none have measured the transcriptome of RA SF neutrophils compared to paired blood neutrophils. In this study we first used RNA-seq to describe the changes that take place when blood neutrophils migrate into RA joints and then validated our bioinformatics predictions using healthy control neutrophils incubated in RA SF *in vitro*. We show that RA SF neutrophils have an altered phenotype, including decreased expression of genes associated with extravasation and migration, increased expression of chemokines, Fc*γ*R1 and MHC II, decreased apoptosis and increased ROS and NET production. All of these altered properties enable SF neutrophils to regulate inflammatory events by attracting and activating innate and adaptive immune cells, including other neutrophils, T cells, NK cells, monocytes, macrophages and dendritic cells within diseased RA joints.

## 2 METHODS

### 2.1 Ethics statement and patient selection

This study was approved by the University of Liverpool Committee on Research Ethics for healthy controls, and NRES Committee North West (Greater Manchester West, UK) for RA patients. All participants gave written, informed consent in accordance with the declaration of Helsinki. All patients fulfilled the ACR 2010 criteria for the diagnosis of RA (Fransen and van Riel, 2005).

### 2.2 Blood and synovial fluid collection

Peripheral blood was drawn into heparinised vacutainers from healthy controls and RA patients. Synovial fluid was aspirated from the knee joint of RA patients (n=3) approximately 1 month prior to the start of the TNFi therapy Etanercept. All patients had a DAS28 score greater than 5.1 at the time of sample collection. Patients had a mean age of 59 years and an average disease duration of 20.6 years; 2 patients were female. Synovial fluid was decanted into universal tubes containing 50*µ*L heparin immediately upon collection and neutrophils were isolated within 1h.

### 2.3 Neutrophil isolation

Neutrophils were isolated from heparinised peripheral blood using Polymorphprep (Axis Shield) as previously described (Wright et al., 2011, 2013). Erythrocytes were lysed using hypotonic lysis with ammonium chloride buffer. Synovial fluid was passed through gauze prior to dilution 1:1 with PBS. Neutrophils were isolated from diluted synovial fluid using Ficoll-Paque (GE Healthcare). Neutrophil purity from both peripheral blood and synovial fluid isolations were > 97% as assessed by cytospin (Supplementary figure 1). Neutrophils accounted for an average of 85.7% leukocytes in whole synovial fluid as assessed by cytospin (Supplementary figure 1). Following isolation, neutrophils were resuspended in RPMI 1640 media (Life Technologies) containing L-glutamine (2mM) and Hepes (25mM) at a concentration of 5×10^6^/mL. Aliquots of synovial fluid were centrifuged at 2000g for 5 min to remove leukocytes. Cell-free synovial fluid was decanted and frozen at -20°C.

### 2.4 RNA extraction

RNA was isolated from 10^7^ neutrophils using Trizol-chloroform (Life Technologies), precipitated in isopropanol and cleaned using the RNeasy kit (Qiagen) including a DNase digestion step. RNA was snap-frozen in liquid nitrogen and stored at - 80°C. Total RNA concentration and integrity were assessed using the Agilent 2100 Bioanalyser RNA Nano chip. RNA integrity (RIN) was ≥ 7.0.

### 2.5 RNA sequencing

Total RNA was enriched for mRNA using poly-A selection. Standard Illumina protocols were used to generate 50 base pair single-end read libraries. Libraries were sequenced on the Illumina HiSeq 2000 platform. Reads were mapped to the human genome (hg38) using TopHat v2.0.4 (Trapnell et al., 2009) and gene expression (RPKM) values were calculated using Cufflinks v2.0.2 (Trapnell et al., 2012). A minimum RPKM threshold of expression of 0.3 was applied to the data in order to minimise the risk of including false positives against discarding true positives from the dataset (Wright et al., 2013; Thomas et al., 2015). Statistical analysis was carried out using Cuffdiff with a 5% false discovery rate (FDR).

### 2.6 Bioinformatics analysis

Bioinformatics analysis was carried out using IPA (Ingenuity Systems, www.ingenuity.com), as previously described, applying a Benjamini-Hochberg correction to p-values for canonical pathway and upstream regulator analysis. Heatmaps were produced using MeV (Saeed et al., 2006) with Euclidean clustering and average linkage. Multivariate partial least squares-discriminant analysis (PLS-DA) was performed on count data, generated using Rsubread (Liao et al., 2019) using mixOmics v6.10.9 (Rohart et al., 2017) working in R v 3.6.3. Modular analysis of gene expression was carried out using the CRAN package tmod (Weiner, 2018) and based on the modular framework for classifying blood genomics studies proposed by Chaussabel and colleagues (Chaussabel et al., 2008).

### 2.7 Measurement of apoptosis

Neutrophils (5×10^5^/mL) were incubated at 37°C in 5% CO_2_ in RPMI 1640 (+Hepes, +L-glutamine) for 18h in the absence or presence of 10% AB serum (Sigma) or 10% cell-free synovial fluid. Following incubation, 2.5×10^4^ cells were diluted in 50*µ*L of HBSS (Life Technologies) containing 0.5*µ*L Annexin V-FITC (Life Technologies), and incubated in the dark at room temperature for 15 min. The total volume was then made up to 500*µ*L with HBSS containing propidium-iodide (PI, 1*µ*g/mL, Sigma Aldrich) before analysis by flow cytometry (>5,000 events analysed) using a Guava EasyCyte flow cytometer.

### 2.8 Measurement of ROS production

Neutrophils (5×10^6^/mL) were incubated with or without TNF*α* (10ng/mL, Merck) for 20 min. Following priming, neutrophils (2.5×10^5^) were diluted in HBSS in the presence of 10*µ*M luminol (Sigma Aldrich). ROS production was stimulated by either f-Met-Leu-Phe (fMLP, 10^−6^M, Sigma Aldrich) or 25% cell-free synovial fluid. Luminol-enhanced chemiluminescence was measured continuously for 60 min on a Tecan plate reader.

### 2.9 Chemotaxis assay

The chemotaxis assay was carried out in 24-well tissue culture plates (coated with 12mg/mL poly-hema to prevent cell adhesion) using hanging chamber inserts with a 3*µ*M porous membrane (Merck), as previously described (Wright et al., 2014a). Media containing fMLP (10^−^8M), interleukin-8 (100ng/mL, Sigma) or 25% (v/v) synovial fluid was added to the bottom chamber. Neutrophils (10^6^/mL) were added to the top chamber and incubated for 90 min at 37°C and 5% CO_2_. The number of migrated cells after 90 min incubation was measured using a Coulter Counter Multisizer-3 (Beckman Coulter).

### 2.10 Assay for neutrophil extracellular trap (NET) production

Neutrophils were seeded (at 2×10^5^ cells/500*µ*L) in RPMI media plus 2% AB serum in a 24-well plate containing poly-L-lysine coated coverslips as previously described (Chapman et al., 2019). Cells were allowed to adhere for 1h prior to stimulation with phorbol 12-myristate 13-acetate (PMA, 50nM, Sigma), A23187 (3.8*µ*M, Sigma) or 10% RA SF. Cells were incubated for a further 4h to allow for NET production. Cells adhered to coverslips were fixed with 4% paraformaldehyde prior to immunofluorescent staining. Briefly, coverslips were removed from the plate and washed with PBS, permeabilised with 0.05% Tween 20 in TBS, fixed with TBS (2% BSA) and then stained for 30 min on drops of TBS (2% BSA) on parafilm stretched across a clean 24-well plate. Primary antibodies used were mouse anti-myeloperoxidase (1:1000, Abcam) and rabbit anti-citrullinated histone H3 (1:250, Abcam). Coverslips were washed three times with TBS prior to secondary antibody staining (anti-rabbit AlexaFluor488, 1:2000, anti-mouse AlexaFluor647, Life Technologies) in TBS (+2% BSA) for 30 min. Coverslips were washed prior to staining with DAPI (1g/mL, Sigma (Sigma). Slides were imaged on an Epifluorescent microscope (Zeiss) using a 40X objective. Images were analysed using ImageJ (Schindelin et al., 2015) and are presented with equal colour balance.

### 2.11 Statistical analysis

Statistical analysis of experimental data was performed using a Student’s t-test or ANOVA in GraphPad Prism (v5) as stated in the text.

## 3 RESULTS

### 3.1 Transcriptomic alternations from blood to synovial fluid

In order to determine the changes in RA neutrophil transcriptome induced by migration from peripheral blood to inflamed, synovial joints, we isolated paired blood and synovial fluid neutrophils from patients with RA (n=3) with active disease (DAS28> 5.1) prior to commencement of biologic therapy with the TNF inhibitor etanercept. Transcriptome sequencing (RNAseq) identified 772 genes that were significantly different between peripheral blood neutrophils and synovial fluid (SF) neutrophils (FDR< 0.05). Of these, 412 genes were significantly higher in blood neutrophils and 347 were higher in synovial fluid neutrophils. Multivariate partial least squares-discriminant analysis (PLS-DA) on the expressed neutrophil transcriptome (∼ 14,000 genes) modelled the difference between PB and SF neutrophil transcriptomes with a ROC AUC = 1 (Figure 1, p< 0.05, 2 components). Modular analysis of the transcriptional networks active in RA blood and SF neutrophils revealed activation of gene modules regulating localisation, cytoskeletal remodelling, and cell signalling (ATP, small GTPases, phosphatidylinositol) in both blood and SF neutrophils. However, signalling in response to MHC, toll-like receptors, inflammasomes and type I interferons were higher in SF neutrophils (Figure 1B). Modules corresponding to integrin signalling and recruitment of neutrophils were higher in blood neutrophils compared to SF.

**Figure 1.**
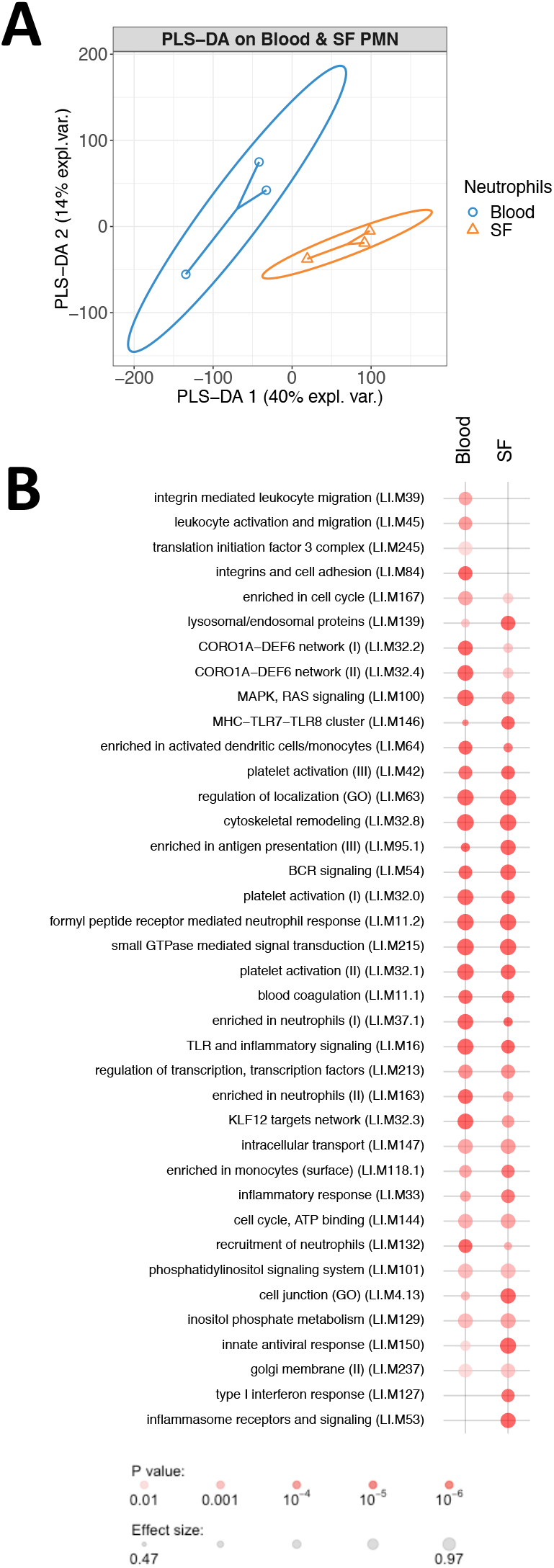
Informatics analysis of blood and synovial fluid (SF) neutrophil transcriptomes. (A) PLS-DA modelling of blood and SF neutrophil transcriptomes using mixOmics. Blood and SF neutrophils were discriminated based on 2 components with an AUC of 1.0 (p< 0.05). (B) Modular analysis of gene expression networks in blood and SF neutrophils using tmod (AUC> 0.8).

In order to determine the signalling pathways most altered in RA neutrophils following migration from peripheral blood to synovial joints, we carried out bioinformatics analysis using Ingenuity Pathway Analysis (IPA) applying a 1.5-fold change in gene expression cut-off. The pathways most significantly up-regulated in SF neutrophils were: Antigen Presentation Pathway (p=10^−8^), Role of NFAT in the regulation of the immune response (p=10^−7^), and Acute phase response signalling (p=10^−6^). EIF2 Signalling (p=10^−8^), STAT3 pathway (p=10^−8^), Leukocyte Extravasation (p=10^−7^) and Granulocyte Adhesion and Diapedesis (p=10^−6^) were the most down-regulated pathways (Figure 2A). Chemokine signalling was up-regulated in RA SF neutrophils (p=10^−4^). Chemokines up-regulated in RA SF (Figure 2B) included those associated with attraction of both innate and adaptive immune cells, including other neutrophils (CXCL8, CXCL1, CXCL2), T cells, monocytes and macrophages, natural killer cells and dendritic cells (CXCL16, CXCL10, CCL2, CCL3, CCL4) (Mantovani et al., 2011; Griffith et al., 2014; Tecchio and Cassatella, 2016). Genes up- and down-regulated genes in the Leukocyte Extravasation Signalling pathway are shown in Figure 2C (p=10^−7^). HIF-1*α* signalling was up-regulated (p=10^−4^), in line with the known hypoxic state of the RA SF joint (Cross et al., 2006), and the HIF-1*α* transcription factor complex was predicted to be activated in RA SF neutrophils (p=5.8×10^−12^).

**Figure 2.**
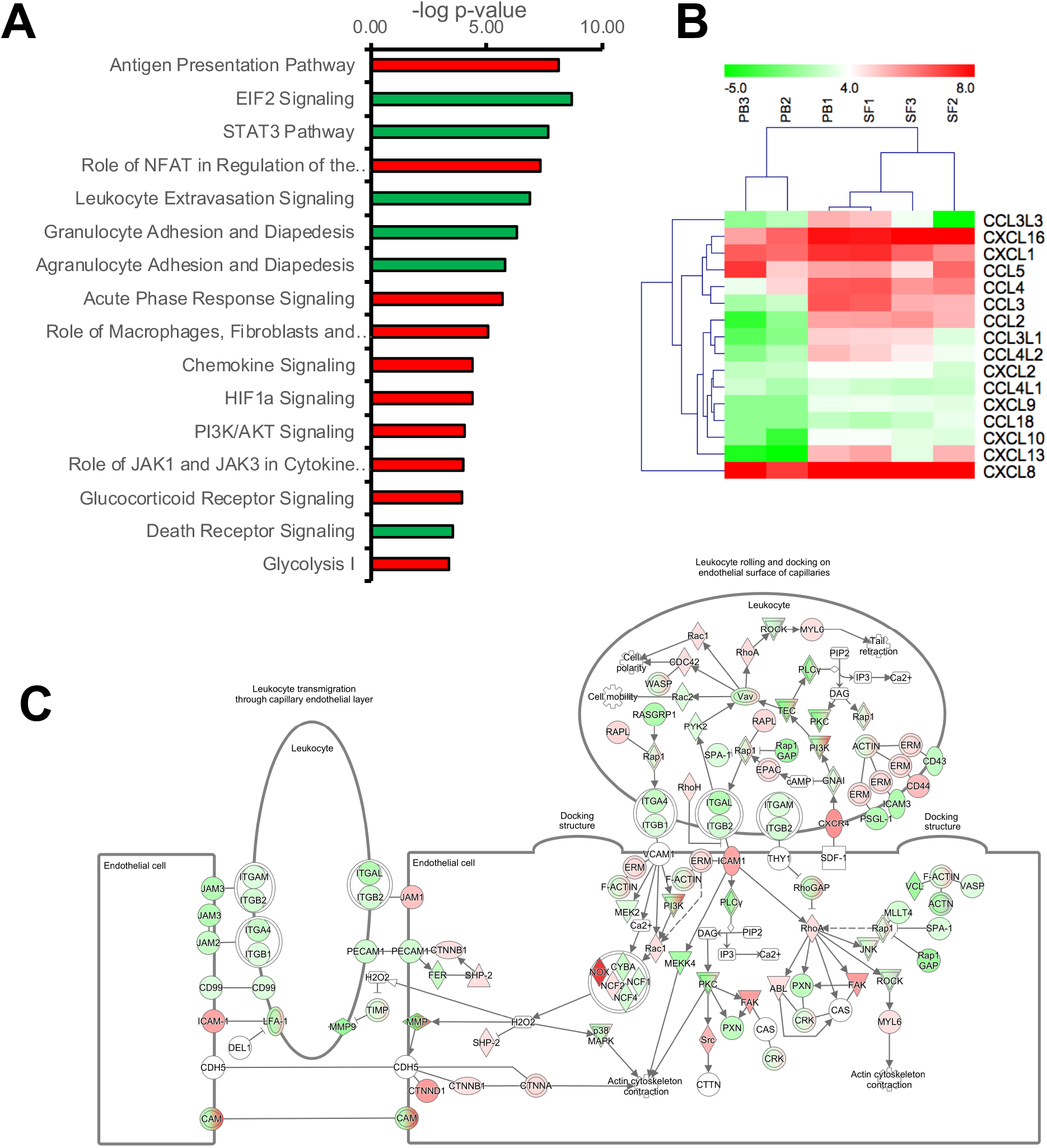
Transcriptomic analysis of peripheral blood (PB) and synovial fluid (SF) neutrophils. (A) IPA signalling pathways predicted to be up-regulated (red) or down-regulated (green) in SF neutrophils. (B) Heatmap showing chemokine gene expression in PB and SF neutrophils. (C) Leukocyte extravasation pathway showing genes up-regulated (red) or down-regulated (green) in SF neutrophils (white = not expressed).

The Antigen Presentation Pathway was predicted to be up-regulated (p=10^−8^), and indeed genes for MHC Class II (HLA-DMA, HLA-DMB, HLA-DQB1, HLA-DRA, HLA-DRB1, HLA-DRB5) were all significantly higher in SF neutrophils in line with previous reports (Cross et al., 2003). Networks of genes regulating the migration and activation of antigen presenting cells were predicted to be up-regulated in SF neutrophils (Figure 3). In particular, these networks were predicted to be regulated by ERK (Figure 3A), MYD88 (Figure 3B) and TICAM1 signalling (Figure 3C). We also noted genes encoding the high affinity Fc*γ*R1 (FCGR1A, FCGR1B, FCGR1C) were significantly higher in RA SF neutrophils (FDR< 0.01), as previously reported (Quayle et al., 1997).

**Figure 3.**
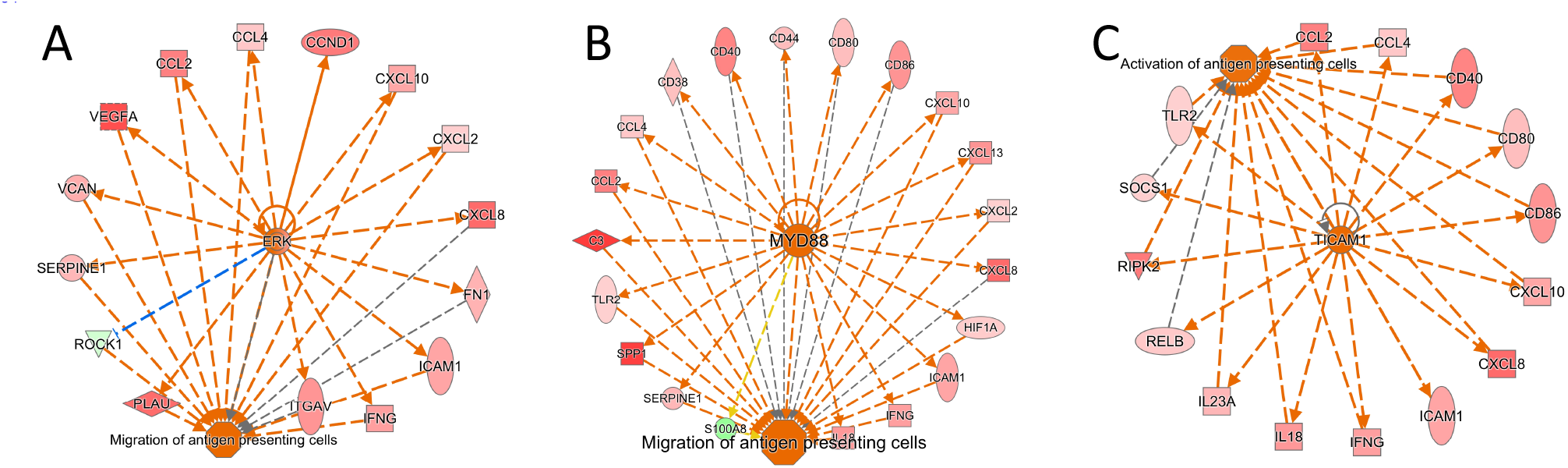
Activation of gene networks in RA SF neutrophils that control migration and activation of antigen presenting cells. These networks are regulated by (A) ERK, (B) MYD88 and (C) TICAM1.

Glycolysis enzymes were highly expressed in RA SF neutrophils (glycolysis pathway activation p=10^−3^). This is in line with our recent metabolomics analysis of RA SF which identified lower levels of substrates for glycolysis in RA SF; this corresponds to higher levels of anaerobic cellular metabolism under hypoxic conditions (Anderson et al., 2018). Lactate dehydrogenase enzymes were also higher in SF neutrophils (LDHA, LDHB; FDR< 0.05). These enzymes catalyse the reduction of pyruvate to lactate producing NAD^+^ and enabling continued energy production under the hypoxic cellular state (Kumar and Dikshit, 2019).

### 3.2 Effect of synovial fluid on neutrophil migration, apoptosis and ROS production

In order to validate the predictions of the IPA analysis, we tested the ability of RA SF to alter neutrophil function. We found that 25% RA SF (3 separate donors, all RF positive) could significantly induce healthy blood neutrophil migration through a 3*µ*M transwell membrane (Figure 4A, ANOVA p< 0.0001) compared to both random migration (Tukey’s post-hoc p< 0.001), and known chemoattractants fMLP (10^−8^M, Tukey’s post-hoc p< 0.001) and interleukin-8 (IL-8, 100ng/mL, Tukey’s post-hoc p< 0.001). We also observed a delay in neutrophil apoptosis (18h) induced by incubating healthy control neutrophils in 10% cell-free SF from the RA neutrophil donors (Figure 4B). This delay in apoptosis was significant for all RA SF compared to untreated (media only) neutrophils (p< 0.05). However, when the RA SF was compared to 10% AB serum, the delay in apoptosis was only significant for SF1 (p< 0.05). Apoptosis was not significantly altered by migration into synovial fluid through a transwell membrane (3*µ*M, data not shown). IPA analysis indicated that this delay in apoptosis was via the up-regulation of the NF*κ*B transcription factor complex (p=2.9×10^−8^) in response to activation of TNF receptors 1 and 2 (TNFR1 signalling p=10^−2^; TNFR2 signalling p=10^−3^) in RA SF neutrophils (Figure 4C).

**Figure 4.**
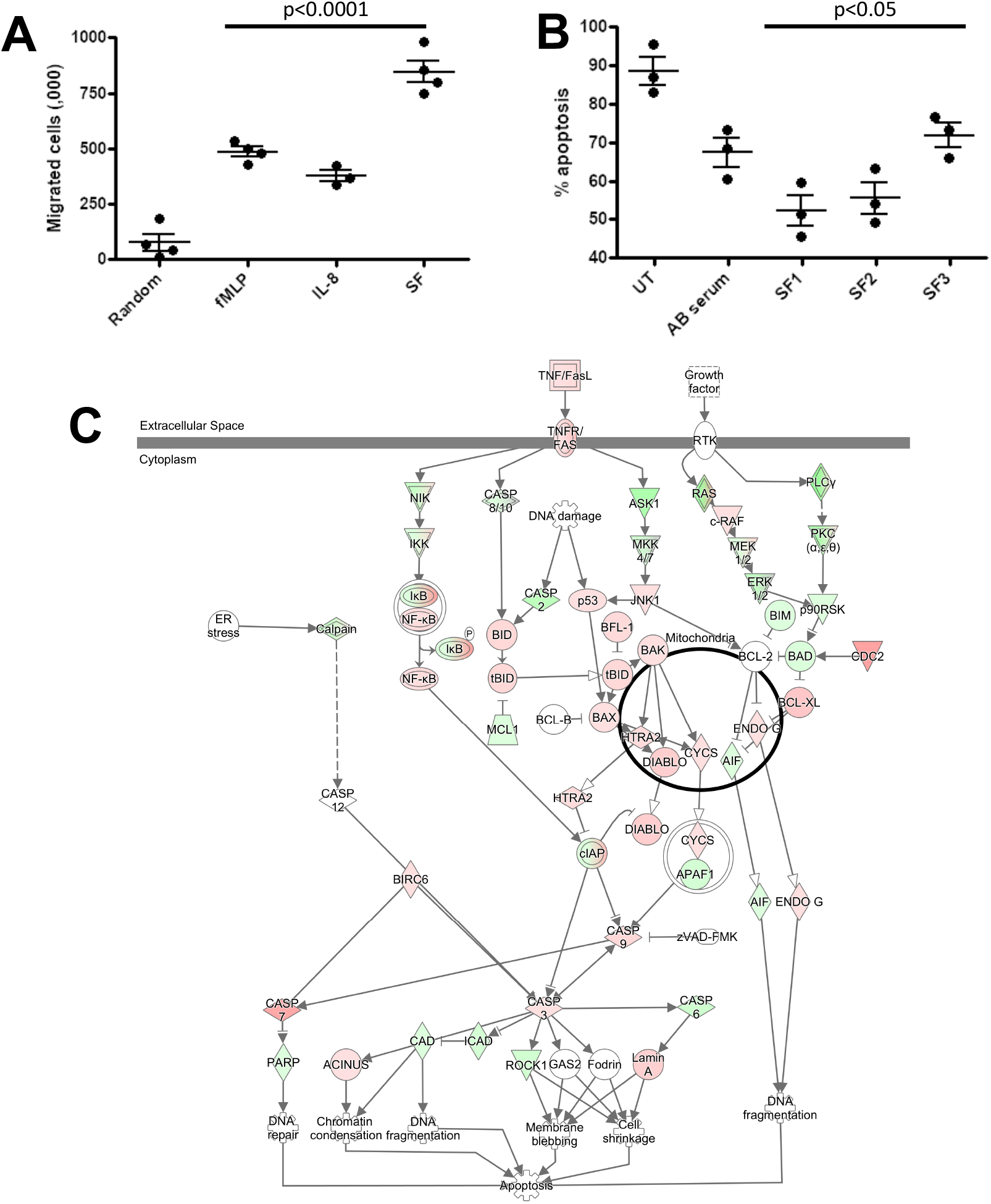
Effect of RA SF on neutrophil migration and apoptosis. (A) Neutrophil chemotaxis was significantly increased towards fMLP, interleukin-8 and 10% RA SF. (B) Neutrophil apoptosis was significantly delayed by RA SF over 18h. (C) IPA apoptosis signalling pathway showing up-regulation (red) and down-regulation (green) of genes associated with regulation of apoptosis in SF neutrophils (white = not expressed).

We next used IPA to predict which cytokines were regulating neutrophil gene expression in RA SF. The major cytokines predicted were interferon-gamma (IFN*γ*), TNF*α*, interleukin-1*β*, interleukin-6 (IL-6) and granulocyte-colony stimulating factor (G-CSF) (Figure 5A, p¡10^−15^). The levels of these cytokines had previously been measured in the donor RA SF as part of a parallel study (Figure 5B) (Wright et al., 2012). Evidence of RA SF neutrophil activation by cytokines can be seen by measuring the respiratory burst in cytokine-primed and unprimed blood and SF neutrophils using luminol-enhanced chemiluminescence. When unprimed blood neutrophils were activated with fMLP, little or no ROS production is observed; this is greatly enhanced by priming for 20 min with TNF*α* (Figure 5C). However, unprimed RA SF neutrophils produced around 2-fold greater ROS compared to unprimed RA blood neutrophils, indicating the SF neutrophils have been primed *in vivo* by synovial cytokines (Figure 5C). Production of reactive oxygen species was predicted by IPA canonical pathway analysis (p=10^−3^, data not shown).

**Figure 5.**
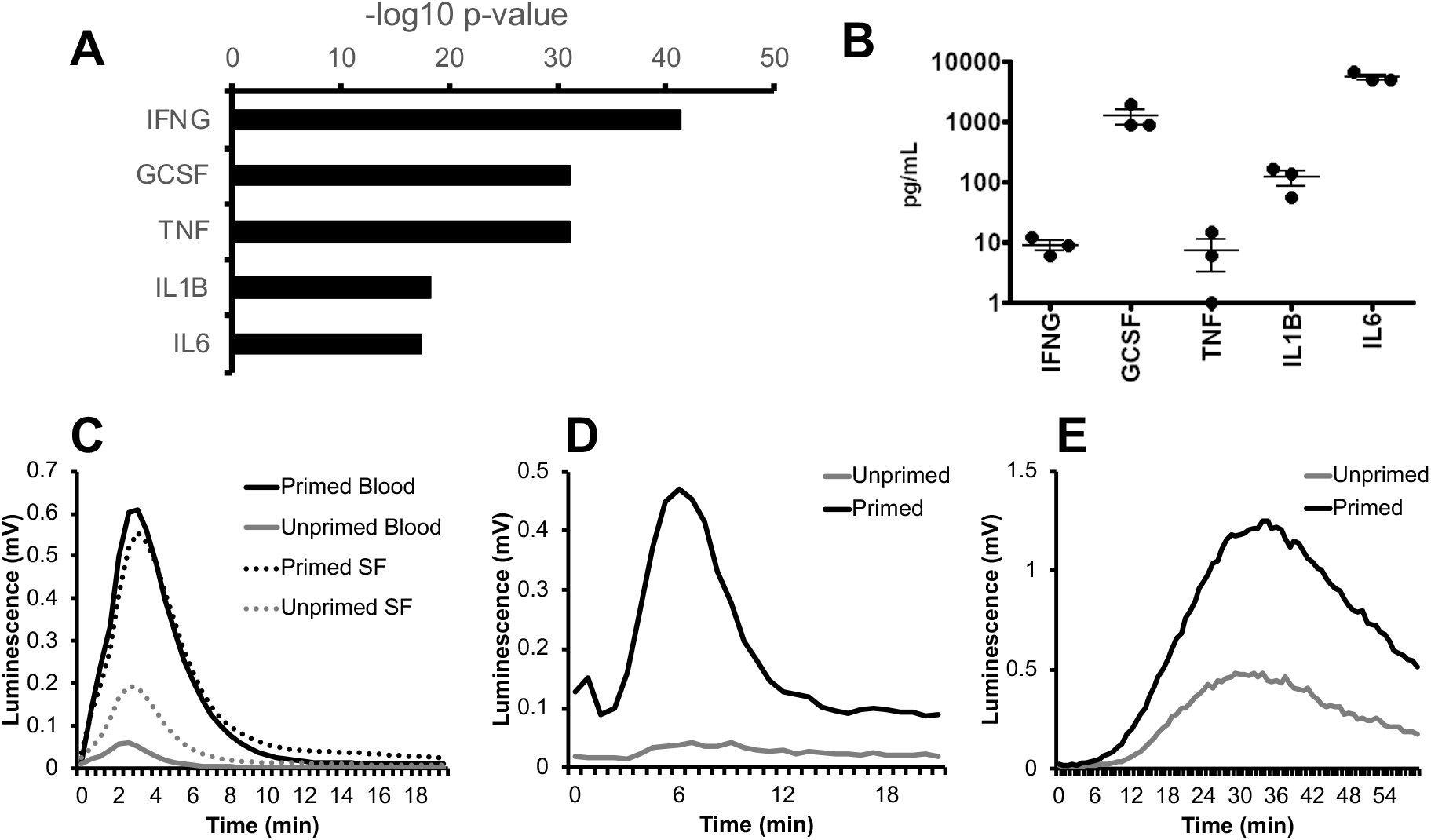
Cytokines in RA SF prime the neutrophil respiratory burst. (A) IPA predicted up-stream cytokine regulators of gene expression in RA SF neutrophils (IFNG, interferon-*γ*; GCSF, granulocyte colony stimulating factor; TNF, tumour necrosis factor *α*; IL1B, interleukin 1*β*; IL6, interleukin 6). (B) Levels of cytokines measured in RA SF from which the neutrophils sequenced by RNA-seq were isolated. (C) ROS production by RA blood and SF neutrophils (with and without cytokine priming) to fMLP. RA SF also activates ROS production in healthy neutrophils. SF containing soluble immune complexes (D) activates ROS production in cytokine primed neutrophils, whereas SF containing insoluble immune complexes (E) activates ROS production in both primed and unprimed neutrophils.

IPA analysis also predicted that RA SF neutrophils had been activated by immunoglobulins (p< 0.01, data not shown); this is likely to be immune complexes such as rheumatoid factor (RF) and/or anti-citrullinated protein antibodies (ACPA). The presence of immune complexes in RA SF can be detected using luminol-enhanced chemiluminescence (Robinson et al., 1992). Soluble immune complexes activate a rapid respiratory burst in cytokine primed neutrophils (Figure 5D) whereas insoluble immune complexes activate a slower and more sustained respiratory burst in both cytokine-primed and unprimed neutrophils (Figure 5E).

### 3.3 Effect of synovial fluid on NET production

NETs are implicated in the pathology of RA by the exposure of antigenic proteins such as citrullinated histones (Wright et al., 2014b; Khandpur et al., 2013). Whilst the exact signalling mechanisms regulating NET production in have yet to be fully elucidated, several signalling pathways that contribute to NET production have been proposed, including: Raf-MEK-ERK, RIPK1-RIPK3-MLKL, AKT, p38-MAPK and cSrc as the four main drivers in NOX2-dependent NETosis (Hakkim et al., 2011; Desai et al., 2016; Amini et al., 2016; Khan and Palaniyar, 2017). Interestingly, all these signalling pathways were predicted by IPA as up-stream regulators of gene expression in RA SF neutrophils (p< 0.01). Signalling networks regulated by AKT, RAF1 and SRC are shown in Figure 6A.

**Figure 6.**
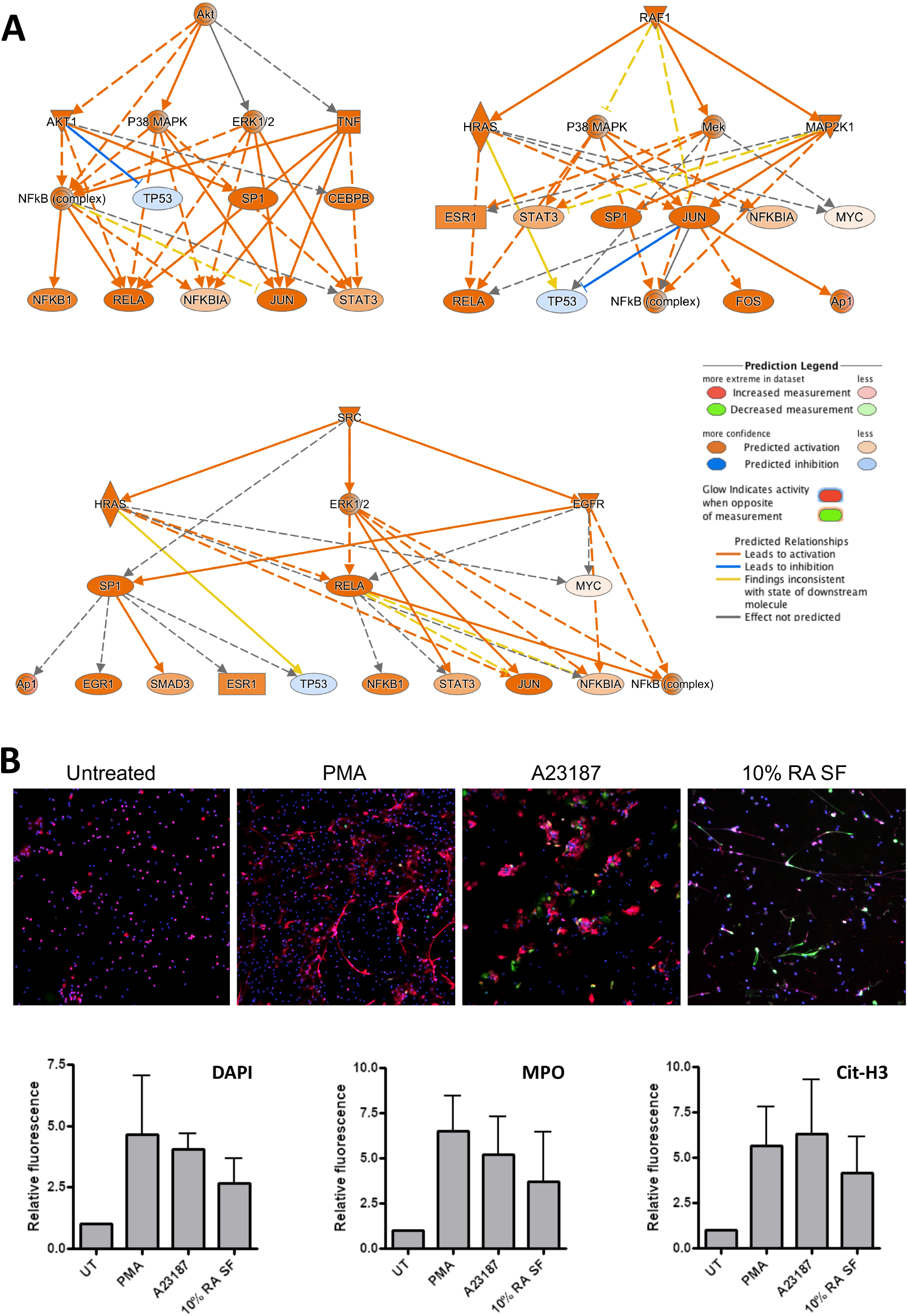
Activation of NET production by RA SF. (A) IPA predicted activation of signalling cascades regulated by AKT, RAF1 and SRC which may regulate NET production. (B) Neutrophils were incubated for 4h with PMA (50nM), A23187 (3.8*µ*M) or 10% RA SF. NET production was increased by RA SF as measured by DAPI staining of extracellular NET DNA (blue), and staining for myeloperoxidase (MPO, red) and citrullinated histone H3 (cit-H3, green) on NET structures.

We therefore tested the ability of RA SF to activate NET production by healthy control neutrophils. We found that 10% RA SF increased NET production by neutrophils to levels similar to that of positive controls PMA and A23187 as shown by the level of extracellular DNA (DAPI), myeloperoxidase (MPO) and citrullinated histone H3 visible on exposed on NET structures (Figure 6B).

## 4 DISCUSSION

In this study we have described for the first time the changes in gene expression that take place in RA neutrophils following migration into inflamed joints. Using RNA-seq we have revealed an activated neutrophil phenotype in RA SF, characterised by increased expression of chemokines, delayed apoptosis and increased activation of kinases and transcription factors which may be implicated in the production of NETs.

Our experiments showed that RA SF decreased the rate of neutrophil apoptosis, which may be attributed to the high levels of inflammatory cytokines present. We previously measured the levels of 13 cytokines in RA SF and found high levels of IL-1*β*, IL-1ra, IL-2, IL-4, IL-8, IL-10, IL-17, IFN-*γ*, G-CSF, GM-CSF and TNF-*α* (Wright et al., 2012). Many of these cytokines have been demonstrated individually to delay neutrophil apoptosis *in vivo*, including G-CSF, GM-CSF, IL-1*β*, TNF*α* and IFN-*γ*, although this is often at concentrations in excess of those found *in vivo* (Cross et al., 2008; Derouet et al., 2004; Moulding et al., 1998; Watson et al., 1998; Maianski et al., 2004; van den Berg et al., 2001; Salamone et al., 2004; Sakamoto et al., 2005). GM-CSF secreted by synovial fibroblasts in response to IL-17 and TNF*α* has been shown to delay neutrophil apoptosis (Parsonage et al., 2008), however a separate study found that apoptosis delay induced by RA SF was not related to the TNF or GM-CSF content, but did correlate with adenosine (Ottonello et al., 2002). Lactoferrin present in RA SF also delays neutrophil apoptosis and may serve as a feed-back anti-apoptotic mechanism for activated neutrophils (Wong et al., 2009). In a separate study, RA SF was shown to be pro-apoptotic in overnight neutrophil cultures, but this effect was reversed when neutrophils were incubated with SF under conditions of hypoxia which more closely model the RA joint (Cross et al., 2006). Relatively few apoptotic neutrophils are present in freshly-isolated RA SF (Raza et al., 2005), and whilst SF is anti-apoptotic, the question remains as to the fate of SF neutrophils. One possibility is that they undergo efferocytosis by synovial macrophages, or another is that they may reverse migrate from the joint back into circulation, as demonstrated in zebrafish models of inflammation (Ellett et al., 2015).

RA neutrophils contain high intracellular levels of citrullinated proteins, mediated through activation of peptidylarginine deiminases (PADs) (Romero et al., 2013). These include known auto-antibody targets: cit-actin, cit-histone H1.3, cit-histone H3, cit-vimentin (Khandpur et al., 2013; Romero et al., 2013). We recently showed that NETs produced by RA neutrophils contain citrullinated proteins, including those proteins previously mentioned as antibody targets: cit-*α*-enolase, cit-histone H2A, cit-histone H4, cit-vimentin, cit-MNDA (Chapman et al., 2019). In our previous study, we also showed that RA NETs contained significantly higher levels of CRISP3 and MMP8, and IL-8 compared to NETs produced by SLE neutrophils. We also reported the presence of methylated and acetylated proteins on RA NETs, particularly acetylated and methylated histones (Chapman et al., 2019). This matches the recent observation that NET debris in RA serum *in vivo* contains high levels of acetylated histones, indicative of NOX2-independent NET production (Pieterse et al., 2018). A recent study reported no association of ACPA with the amount of NET material in RA serum (de Bont et al., 2020). Interestingly, antibodies to NET material (ANETA) were present in ACPA negative patients as well as ACPA positive, although ANETA were significantly higher in those patients seropositive for RF (de Bont et al., 2020).

The RAGE-TLR9 pathway plays a key role in both the internalisation and presentation of citrullinated NET peptides on MHC Class II in fibroblast-like synoviocytes (FLS). This leads to the development of ACPA specific to the citrullinated NET peptides and cartilage damage in mouse models of RA (Carmona-Rivera et al., 2017). A role for NET-derived elastase in cartilage destruction has also been proposed, whereby elastase contained in NET material disrupts the cartilage matrix and induced the release of PAD2 by fibroblast-like synoviocytes (FLS) leading to citrullination of cartilage fragments (Carmona-Rivera et al., 2020). These citrullinated cartilage fragments are subsequently internalised and presented by FLS and macrophage to antigen specific T-cells leading to the development of auto-immunity and ACPA in HLA-DRB1*04:01 transgenic mice (Carmona-Rivera et al., 2020).

It has previously been shown that levels of ACPA in RA SF correlate with neutrophil numbers and severe disease activity, and that SFs with high ACPA titres induce high levels of ROS and NET production (Gorlino et al., 2018). A separate study demonstrated that depletion of ACPA from RA SF inhibits NET production *in vitro* (Sur Chowdhury et al., 2014). A role for IgA immune complexes in RA SF has also been proposed, with SF rich in IgA inducing NET and ROS production and release of lactoferrin by healthy control neutrophils in a mechanism that was inhibited by blockade of the Fc*α*RI receptor (Aleyd et al., 2016). RA SF also contains high levels of extracellular DNA, concentrations of which correspond to neutrophil counts and PAD activity, and which have been attributed to NETs (Spengler et al., 2015). Our recent proteomics study identified both PAD2 and PAD4 in RA NETs (Chapman et al., 2019). PAD enzymes have also been identified by IHC in synovial biopsies, localised with MPO in necrotic areas of synovial tissue (Turunen et al., 2016) that contain large areas of citrullinated and hypercitrullinated proteins, possibly indicating the presence of NET structures within synovial tissue.

Whilst the exact signalling mechanism for NET production remains unclear, at least two modes of NET production (NETosis) have been described: NOX2-dependent and NOX2-independent. NOX2-dependent NETosis occurs via activation of the NADPH-oxidase (NOX2) and production of ROS. This causes increased intracellular membrane permeability, elastase release into the nucleus and degradation of histones leading to chromatin decondensation and NET release (Papayannopoulos et al., 2010). NOX-2 independent NETosis does not require the production of ROS by the NADPH oxidase. In this case, mitochondrial ROS combine with increased intracellular calcium levels to activate PADs leading to hypercitrullination of histones, chromatin decondensation and NET release (Neeli et al., 2008; Douda et al., 2015). Signalling pathways including Raf-MEK-ERK, RIPK1-RIPK3-MLKL, AKT, p38-MAPK and cSrc have been identified as some of the drivers of NETosis (Hakkim et al., 2011; Desai et al., 2016; Amini et al., 2016; Khan and Palaniyar, 2017). In our study, all of these signalling pathways were predicted to be activated in RA SF neutrophils. Interestingly, all of the kinase activation networks predicted also included downstream activation of NF-*κ*B. This important signalling pathway, activated by cytokines such as TNF*α* and IL-1*β* has not yet been implicated in NET production and would be an interesting candidate for future investigation.

Our RNA-seq analysis identified increased expression of MHC Class II genes in RA SF neutrophils. We have previously shown that whilst healthy control neutrophils do not express MHC Class II, RA blood neutrophils express MHC Class II RNA, and RA SF neutrophils expression both RNA and MHC Class II protein, although the latter was contained within intra-cellular pools which were mobilised to the plasma membrane following overnight incubation (Cross et al., 2003). Expression of co-stimulator molecules CD80 and CD86 was only detected at very low levels. However, SF neutrophils were able to stimulate CD4^+^ T-cell proliferation via a mechanism that could be inhibited by an anti-MHC Class II antibody (Cross et al., 2003). We also detected increased expression of a number of chemokines in RA SF, including CCL3, CCL4, CCL10,, CXCL16, CXCL2, CXCL8. These chemokines play a key role in regulating the inflammatory response in the joint, not only through recruitment of other neutrophils (CXCL1, CXCL2, CXCL8), but also through the recruitment and activation of both innate and adaptive immune cells (Mantovani et al., 2011; Griffith et al., 2014; Tecchio and Cassatella, 2016) as summarised in Figure 7. This increased production of chemokines within the joint, coupled with a down-regulation of adhesion receptors, suggests that RA SF neutrophils become resident within the joint to drive further inflammation through recruitment and activation of other immune cells.

**Figure 7.**
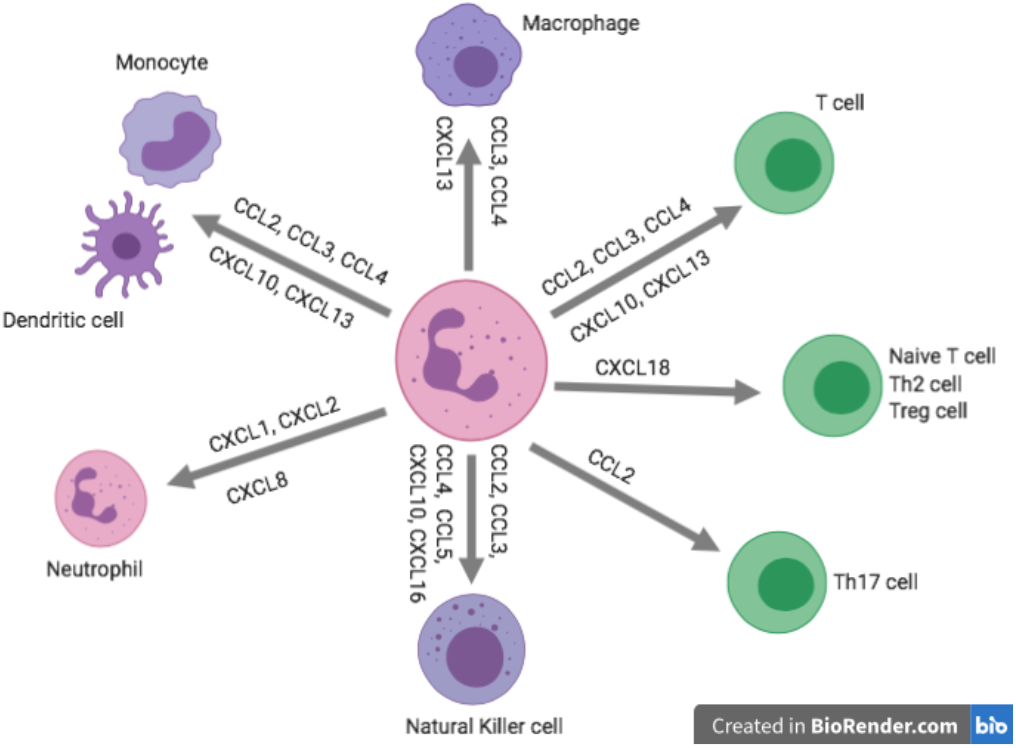
Summary of the role of inflammatory role of chemokines expressed by RA SF neutrophils.

Whilst a predominantly pro-inflammatory role for SF neutrophils has been proposed in our study, several studies have suggested a pro-resolving role for neutrophils in inflammatory arthritis (Headland and Norling, 2015). For example, neutrophil-derived microvesicles may be anti-inflammatory in animal models of inflammatory arthritis through expression of the pro-resolving protein annexin-A1 (Headland et al., 2015). Annexin-A1 is released from neutrophil granules following extravasation and is found in synovial fluid. This protein has many pro-resolving functions, including promoting apoptosis and decreasing neutrophil:endothelial cell adhesion and extravasation (Headland and Norling, 2015; Perretti and Solito, 2004). Indeed, annexin-A1 knock out mice suffer from exacerbated synovial inflammation and cytokine production in a methylated BSA-induced model of joint inflammation (Yang et al., 2004). Nanoparticles coated in neutrophil membranes decrease cytokine production, suppress synovial inflammation and prevent joint damage in both collagen-induced arthritis and a human transgenic mouse model of arthritis (Zhang et al., 2018). In gout, aggregated NETs degrade a number of pro-inflammatory cytokines, including IL-6, TNF*α* and IL-1*β*, and promote resolution of inflammation (Schauer et al., 2014) and in an infection model, neutrophil granule proteins degraded inflammatory proteins, including TNF*α* and IL-1*β* (Gresnigt et al., 2012).

Our findings provide novel insight into the multitude of ways that synovial neutrophils drive chronic inflammation in RA. This raises the possibility that aberrant neutrophil activation may be a target of future therapeutics in this chronic and life-limiting condition. A number of new therapies which directly or indirectly target neutrophil function have been proposed or are in clinical trial (Nemeth et al., 2020). Inhibitors of CXCR1/CXCR2, the receptor for CXCL8 (interleukin-8), have been demonstrated to reduce neutrophil adhesion and recruitment to synovial joints in murine models of inflammatory arthritis (Cunha et al., 2008), an effect which was associated with lower TNF*α* levels and disease activity (Coelho et al., 2008). JAK inhibitors including tofacitinib and baricitinib, which target signalling down-stream of cytokine receptors including IL-6, interferon-*α* and -*γ*, and GM-CSF receptors, are clinically effective in treating RA (Burmester et al., 2013; Taylor et al., 2017) although many patients report transient neutropenia and increased infections. These drugs have been shown to inhibit cytokine signalling in neutrophils, including inhibition of migration and ROS production by RA neutrophils (Mitchell et al., 2017). The anti-GM-CSF therapy mavrilimumab was effective in decreasing RA disease activity in a Phase 2b clinical trial, with over 70% of RA patients achieving an ACR20 improvement in the group receiving the highest dosing regime (Burmester et al., 2017). Anti-G-CSF therapy prevents neutrophil migration into joints, suppressed cytokine production and halting the progression of murine arthritis (Campbell et al., 2016). Finally, a number of novel therapeutics which target neutrophil proteases, including MPO and elastase, have been effective in reducing neutrophil-driven inflammation in animal models of inflammatory disease (Zheng et al., 2015) and human respiratory disease (Stockley et al., 2013) respectively. These inhibitor drugs have potential to inhibit NET production and damage to cartilage associated with inflammation in RA synovial joints, and thus may be potential therapeutics for repurposing to target neutrophil-driven RA joint inflammation.

In conclusion, we have used RNA-seq and experimental analysis of paired blood and SF RA neutrophils to describe a pro-inflammatory SF neutrophil phenotype which includes delayed apoptosis, production of ROS, and release of NETs. RA SF neutrophils down-regulate adhesion molecules to become resident in the joint and drive inflammation via increased production of chemokines that attract and activate both innate and adaptive immune cells. We propose this altered neutrophil phenotype contributes to the pro-inflammatory nature of active RA and explains the role of neutrophils in the pathogenesis of the disease.

## Data Availability

The datasets generated during and/or analysed during the current study are available from the corresponding author on reasonable request.

## AUTHOR CONTRIBUTIONS

HLW designed the research, carried out the experiments, analysed the data, and wrote the manuscript. ML carried out the experiments and analysed the data. EAC carried out the experiments, analysed the data and revised the manuscript. RJM revised the manuscript. SWE analysed the data and revised the manuscript.

## ACKNOWLEDGMENTS

This work was undertaken on Barkla, part of the High Performance Computing facilities at the University of Liverpool, UK. We also thank Dr RC Bucknall for help with collection of synovial fluid.

